# Evaluation Methods for T-association of a Surrogate Endpoint

**DOI:** 10.1101/2025.08.28.25334653

**Authors:** Jo-Ying Hung, Chih-Yuan Hsu, Pei-Fang Su, Yu Shyr

**Author notes:** These authors contributed equally to this work (Jo-Ying Hung, Chih-Yuan Hsu).

## Abstract

A surrogate endpoint is a biomarker that is reasonably likely to predict clinical benefit and is used as a substitute for a direct measure of clinical benefit under the Food and Drug Administration (FDA) Accelerated Approval pathway. According to FDA guidelines, a valid surrogate endpoint must meet two associations: I-association (the association between the surrogate and true endpoints, such as disease response and overall survival) and T-association (the association between treatment effects on both endpoints, such as odds ratio and hazard ratio). I-association is commonly evaluated, but T-association is often overlooked due to the lack of appropriate statistical methods. Failure to satisfy T-association precludes a biomarker from supporting accelerated approval. To address this gap, we propose a new method to rigorously assess T-association in accordance with FDA guidelines. This method assumes that treatment effects on the surrogate and true endpoints follow a bivariate normal distribution, accounting for both within-study and between-study variances. The key evaluation metric is the correlation coefficient, which quantifies the relationship between treatment effects on both endpoints. Model parameters, including this correlation, are estimated using maximum likelihood, restricted maximum likelihood, and a Bayesian approach. We demonstrate the method using both simulated and real-world data. The method will serve as the statistical foundation that aligns with FDA guidelines and supports future accelerated approvals. The R package to implement the proposed method is available at https://github.com/jybelindahung/T-association.

## 1 Introduction

To support effectiveness claims in new drugs applications to the Food and Drug Administration (FDA), appropriate endpoints must be selected for clinical trials. Endpoints used to assess clinical benefit include both clinical endpoints and surrogate endpoints. Clinical endpoints directly reflect how a patient feels, functions, or survives, and are considered the most reliable measure of treatment benefits. In contrast, surrogate endpoints are biomarkers expected to predict clinical benefit and serve as substitutes for clinical endpoints. Because they can be measured earlier or more easily, surrogate endpoints can potentially shorten trial duration and improve compliance, cost-effectiveness, and ethical considerations.

The FDA provides two approval pathways for new drugs: traditional approval and accelerated approval. Traditional approval requires direct evidence of clinical benefit, as demonstrated by clinical endpoints or validated surrogate endpoints. Accelerated approval, in contrast, may be granted on the basis of intermediate clinical endpoints or surrogate endpoints that are reasonably likely to predict clinical benefit [1]. It allows approval of drugs for serious or life-threatening conditions that address unmet medical need. Since the first establishment of accelerated approval in 1992, the accelerated approval pathway has been increasingly utilized. Figure 1 illustrates the growing number of accelerated approval drugs in recent years based on an FDA report [2]. The trend reflects the expanding role of accelerated approval in drug development. To ensure regulatory rigor under this pathway, the validation of whether a biomarker is a surrogate endpoint is an important issue.

**Figure 1.**
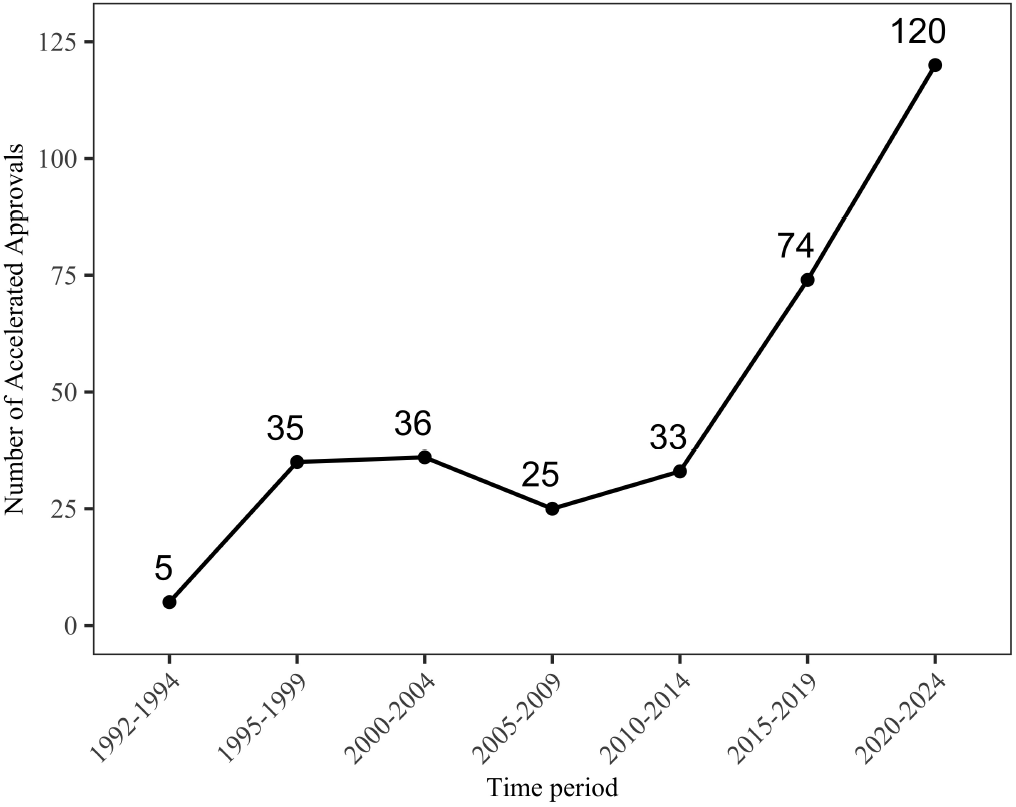
Number of Drugs Granted Accelerated Approval by Year Range (1992–2024).

To determine whether a biomarker can be considered a surrogate endpoint, FDA has outlined statistical principles for validation, including individual-level association (I-association) and trial-level association (T-association)[3]. I-association assesses whether the biomarker correlates with the true clinical endpoint within individuals, while T- association refers to whether the treatment effect on the biomarker is associated with the treatment effect on the true endpoint across trials. For instance, in the aforementioned FDA guideline [3], it is discussed that although minimal residual disease (a biomarker that measures tumor burden) is widely used to reflect a patient’s response to treatment, validating it as a surrogate endpoint requires evidence of both I-association and T-association.

The definitions of I-association and T-association were first proposed by Buyse et al. [4]. They defined coefficients of determination estimated within trials and across trials, and proposed that a biomarker can be considered a valid surrogate only if it demonstrates both individual- and trial-level validity. Despite the rigorous definition, many applied studies still rely primarily on I-association. For instance, Scher et al. [5] evaluated the individual-level surrogacy of circulating tumor cells for survival in prostate cancer; Fokas et al. [6] investigated the individual-level surrogacy of the neoadjuvant rectal score for disease-free survival in rectal cancer; and Yee et al. [7] attempted to identify pathologic complete response as a surrogate for survival endpoints in breast cancer based solely on strong I-association. These approaches do not follow FDA guidelines and thus the biomarkers are not considered valid surrogate endpoints.

A possible reason for the limited evaluation of T-association is that existing statistical methods may be complex and difficult to interpret [4, 8]. As a result, some recent work has adopted simpler approaches; for example, Ye et al. [9] evaluated T-association using the Spearman correlation between estimated treatment effects, an approach that does not account for the within-study uncertainty of the treatment effect estimates. To facilitate a more rigorous evaluation of T-association, we propose a new method based on the bivariate random-effects meta-analysis (BRMA) model [10], which can be used to synthesize two correlated endpoints. The model jointly accounts for both within-study and between-study variances in the estimated treatment effects on the biomarker and the true endpoint, allowing individual weighting of each study. Both frequentist and Bayesian frameworks are applied for estimation and inference. By quantifying T- association through the estimated correlation, our approach provides a practical and interpretable framework that aligns with FDA guidance for surrogate endpoint validation.

This article is organized as follows. Section 2 introduces the proposed model and details both frequentist and Bayesian approaches for parameter estimation. Section 3 presents simulation studies and two real-world applications. Finally, Section 4 discusses the findings.

## 2 Methods

### 2.1 Model Setup

For each study *i* = 1, …, *n*, we let *y*_*i*1_ and *y*_*i*2_ denote the statistics quantifying the treatment effect on the biomarker of interest and the true endpoint, respectively. For example, *y*_*i*1_ and *y*_*i*2_ are the estimates of log(OR) and log(HR), which reflect the treatment effects on dichotomous biomarkers and time-to-event endpoints. Here, OR and HR refer to odds ratio and hazard ratio, respectively. When the number of participants within study *i* is large, both *y*_*i*1_ and *y*_*i*2_ are typically assumed to approximate a normal distribution.

To evaluate T-association, we apply the BRMA model proposed by Riley et al. [10] to synthesize the two treatment effect estimates. The BRMA model can be expressed as follows:

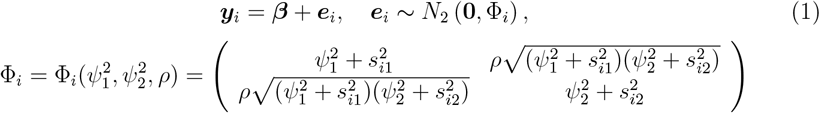

where ***y***_*i*_ = (*y*_*i*1_, *y*_*i*2_)^*′*^ and ***β*** = (*β*_1_, *β*_2_)^*′*^, in which the prime symbol (·)^*′*^ denotes the transpose of a vector or matrix. The parameters *β*_1_ and *β*_2_ represent the mean treatment effects for the biomarker and the true endpoint, respectively. The error term ***e***_*i*_ is assumed to follow a normal distribution with the covariance Φ_*i*_. Within Φ_*i*_, the parameters 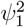 and 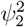 capture the between-study variances, while 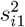 and 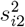 are the known within-study variances, which are typically reported in study *i*. The correlation parameter *ρ* is the key evaluation metric of T-association.

This model offers two advantages. First, it accounts for between- and within-study variances, allowing each study to be appropriately weighted based on its precision through 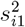 and 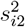. Second, it captures the overall correlation between treatment effects using a single parameter *ρ*, rather than decomposing it into between- and within-study correlations, which is practical given that within-study correlations are rarely available. In the following sections, we present frequentist and Bayesian approaches for obtaining point and interval estimates for the parameters of the BRMA model.

### 2.2 Frequentist Approach

Under the frequentist framework, maximum likelihood (ML) and restricted maximum likelihood (REML) methods are commonly used to estimate model parameters. In the following, we detail the procedures for obtaining ML and REML estimates, along with their associated confidence intervals (CI). Let 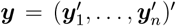 and 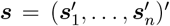, where ***s***_*i*_ = (*s*_*i*1_, *s*_*i*2_)^*′*^, *i* = 1, …, *n*.

#### Maximum likelihood

Let 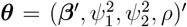, where ***β*** = (*β*_1_, *β*_2_)^*′*^. Given the observed data ***y*** and ***s***, the full likelihood function can be expressed as:

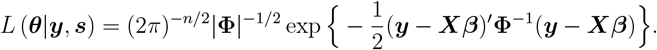

The corresponding log-likelihood function is:

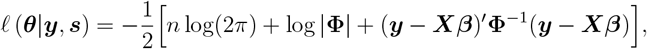

where ***X*** = (**I**_2_, …, **I**_2_)^*′*^ with **I**_2_ denoting the 2 by 2 identity matrix, and **Φ** is a block-diagonal matrix with each study’s covariance Φ_*i*_ along the diagonal, i.e., **Φ** = diag(Φ_1_, …, Φ_*n*_). The ML estimate of 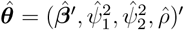 is obtained by solving ***U*** (***θ***) = *∂ℓ* (***θ***|***y, s***) */∂****θ*** = **0**. The detailed expression for the score function ***U*** (***θ***) is provided in the Supplementary Materials. Since no closed-form solution exists for the equation, the ML estimate is obtained iteratively as follows:

##### Step 1

Set the initial parameter value as 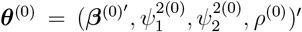. To ensure numerical stability and enforce parameter constraints (positive variances and |*ρ*| *<* 1), the variance components are reparameterized on the log scale, and the correlation is reparameterized using the Fisher *z*-transformation, which is defined as

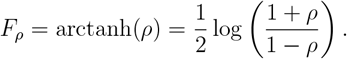

##### Step 2

Solve the system of score equations ***U*** (***θ***) = **0** using the Newton–Raphson algorithm, starting from the initial value ***θ***^(0)^. Continue the iterations, indexed by *m*, until convergence, which is defined by ∥***θ***^(*m*)^ − ***θ***^(*m*−1)^∥ *< ε* for a small tolerance *ε >* 0. In our implementation, the score equations are solved by using the `nleqslv` function from the `nleqslv` package in `R`.

#### Restricted maximum likelihood

Under the REML framework, the restricted log-likelihood function is obtained from integrating out ***β*** from *L*(***θ***|***y, s***), which yields unbiased estimates for variance components.

Let 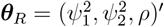, then the restricted log-likelihood is given by:

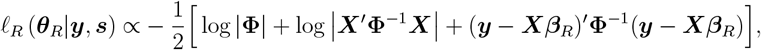

where ***X*** and **Φ** are defined as in the above section, and ***β***_*R*_ = (***X***^*′*^**Φ**^−1^***X***)^−1^***X***^*′*^**Φ**^−1^***y***. Similar to the ML approach, we obtain the REML estimate of 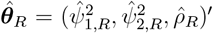 by solving ***U***_*R*_(***θ***_*R*_) = *∂ℓ*_*R*_ (***θ***_*R*_|***y, s***) */∂****θ***_*R*_ = **0** using the Newton-Raphson algorithm. The explicit expression for the score function ***U***_*R*_(***θ***_*R*_) is detailed in the Supplemental Materials. Finally, once 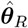 is obtained, the estimate for ***β*** is given by 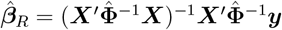, where 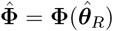.

#### Confidence intervals

To quantify the uncertainty of the estimators and facilitate statistical inference, we construct CIs for all model parameters. Under standard regularity conditions, the ML and REML estimators are asymptotically normal, with variances estimated from the inverses of their observed information matrices, denoted by ℐ and ℐ_*R*_:

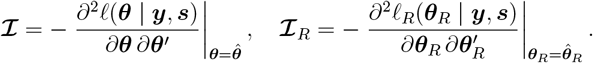

For the REML estimator 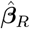, the variance is estimated by 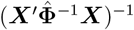. Detailed expressions for the observed information matrices are provided in the Supplemental Materials. CIs are then derived based on the asymptotic normality of the estimators. As mentioned previously, the variance and correlation parameters were reparameterized when solving the score functions. Accordingly, CIs for ***β*** are computed on the original scale, while CIs for the variance components 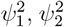, and the correlation *ρ* are first formed on transformed scales to respect parameter constraints and then back-transformed. Specifically, CIs for variance components are computed on the log scale:

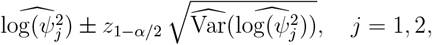

and then exponentiated to return to the original scale, where *z*_1−*α/*2_ is the standard normal quantile and *α* is the pre-specified significance level. CI for the correlation is computed on the Fisher-transformed scale:

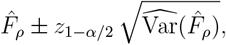

and back-transformed using the hyperbolic tangent. The variance estimates for the estimates of the transformed parameters under ML and REML: 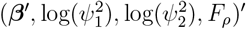 and 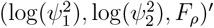 are obtained from (***J*** ^*′*^ℐ***J***)^−1^ and 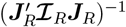, respectively, where 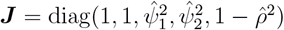 and 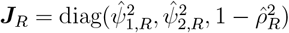.

In addition to these large-sample normal-based intervals, we also apply a parametric bootstrap procedure to construct CIs for the model parameters under both ML and REML frameworks.

### 2.3 Bayesian Approach

In real-world applications, the number of studies available to evaluate T-association may be limited. As a more robust alternative in limited study scenarios, we adopt Bayesian methods. Given the observed data (***y, s***) and a prior distribution *π*(***θ***) for the parameters, the posterior distribution of ***θ*** is expressed as

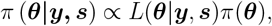

where the full likelihood function *L*(***θ***|***y, s***) is defined in the maximum likelihood section. In this study, we consider the following priors:

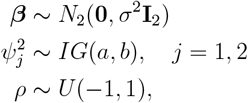

where *IG*(*a, b*) is the inverse-gamma distribution with shape parameter *a* and scale parameter *b*, and *U* (−1, 1) is the uniform distribution over (−1, 1). To ensure weak informativeness, we choose a large value for *σ*^2^ and small values for both *a* and *b*. The uniform prior on *ρ* is commonly used to reflect a flat, noninformative distribution over the feasible correlation range.

Although the inverse-gamma priors with small shape and scale parameters are often used as noninformative priors for variance components, Gelman [11] criticized them for placing excessive mass near zero. As an alternative, they suggested a half-*t* prior, a folded Student’s-*t* distribution truncated to be positive, for *ψ*_*j*_. Gelman [11] showed that this prior provides better regularization and more stable inference for variance parameters. Accordingly, we also consider:

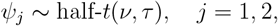

where half-*t*(*ν, τ*) denotes a half-*t* distribution with degrees of freedom *ν* and scale parameter *τ*. For the correlation, in addition to the uniform prior, we also consider a Normal prior on the Fisher-transformed correlation:

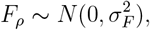

which induces a mild shrinking effect toward zero and serves as a weakly informative prior when 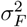 is appropriately chosen. Thus, four prior combinations are provided: 2 priors for variance components and 2 priors for correlation.

Since the resulting posterior distributions do not have closed-form expressions, we employ Markov Chain Monte Carlo (MCMC) to sample from the implicit posterior distributions. Specifically, we adopt Hamiltonian Monte Carlo (HMC) [12], a modern and efficient technique that leverages gradient information from the posterior to propose distant, high-probability values. Compared to traditional random-walk-based methods like Metropolis-Hastings, HMC significantly improves sampling efficiency, especially in settings where parameters exhibit strong correlations [13].

We implement HMC using the `cmdstan model` function from the `cmdstanr` package in `R` [14]. The function uses the No-U-Turn Sampler, an adaptive variant of HMC that automatically tunes the trajectory length to improve sampling efficiency [13]. The posterior distribution is constructed using four parallel Markov chains. Each chain is run for 1,000 warm-up iterations followed by 1,000 sampling iterations, yielding a total of 4,000 post–warm-up draws. Posterior means of the parameters ***θ***, including the key T- association metric *ρ*, are used as point estimates. The 100(1 − *α*)% credible intervals (CrI) are constructed using the *α/*2 and 1 − *α/*2 quantiles of the corresponding marginal posterior distributions, where *α* is the pre-specified credible level.

## 3 Numerical Studies

### 3.1 Simulation Studies

We conducted simulation studies to assess the performance of the proposed methods. In the simulations, the treatment effects for the biomarker and the true endpoint were generated from model (1) with parameters ***β***^*′*^ = (0.6, −0.2) and 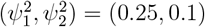. These values were selected to reflect realistic treatment effect magnitudes and between-trial variability observed in real data [9]. Moreover, to investigate the impact of correlation, the parameter *ρ* was set to 0.1, 0.4, and 0.7, corresponding to low, moderate, and high correlations. Within-study standard errors *s*_*i*1_ and *s*_*i*2_, assumed to be known, were generated from a uniform distribution *U* (0.05, 0.5). The number of trials we consider include: *n* = 6, 10, and 15. For each scenario, 1,000 simulation replicates were performed, and estimation results from both frequentist and Bayesian approaches are reported.

#### Frequentist approach

Parameter estimates were obtained using both ML and REML methods. Estimation performance was evaluated based on empirical bias (Bias), empirical standard error (SE_*em*_) calculated from 1,000 simulation replicates, the average theoretical standard error (SE_*th*_) across 1,000 replicates, and the coverage probability of the 95% CI (CP_*th*_) based on the theoretical standard error. In addition, the coverage probability of the 95% CI obtained via parametric bootstrap (CP_*b*_) was calculated using 1,000 bootstrap samples. Results are presented in Table 1.

**Table 1.** Simulation results under the frequentist framework across different numbers of trials (*n*) and true correlations (*ρ*_true_). The significance level *α* was set to 0.05. “Par” stands for parameter.

| $n$ | $\rho_{\text{true}}$ | Par | ML | | | | | REML | | | | |
| --- | --- | --- | --- | --- | --- | --- | --- | --- | --- | --- | --- | --- |
|  |  |  | Bias | SE<sub>th</sub> | SE<sub>em</sub> | CP<sub>th</sub> | CP<sub>b</sub> | Bias | SE<sub>th</sub> | SE<sub>em</sub> | CP<sub>th</sub> | CP<sub>b</sub> |
| 15 | 0.1 | $\rho$ | -.003 | .239 | .266 | 91.8 | 93.7 | -.003 | .247 | .266 | 93.3 | 93.9 |
| | | $\beta_1$ | .000 | .141 | .149 | 91.6 | 91.3 | .000 | .145 | .148 | 92.6 | 92.1 |
| | | $\beta_2$ | -.001 | .103 | .105 | 93.4 | 92.6 | -.001 | .106 | .104 | 94.2 | 94.2 |
| | | $\psi_1^2$ | -.021 | .111 | .114 | 94.7 | 84.4 | .002 | .124 | .122 | 96.3 | 90.7 |
| | | $\psi_2^2$ | -.004 | .057 | .054 | 98.0 | 89.1 | .009 | .065 | .058 | 97.8 | 94.4 |
| | 0.4 | $\rho$ | .001 | .203 | .230 | 91.9 | 93.5 | .000 | .211 | .230 | 93.3 | 93.6 |
| | | $\beta_1$ | -.002 | .140 | .148 | 91.5 | 90.3 | -.002 | .144 | .148 | 92.4 | 92.6 |
| | | $\beta_2$ | .000 | .103 | .105 | 93.2 | 92.3 | .000 | .105 | .104 | 94.0 | 93.9 |
| | | $\psi_1^2$ | -.021 | .111 | .114 | 94.8 | 85.2 | .002 | .124 | .122 | 95.9 | 90.5 |
| | | $\psi_2^2$ | -.004 | .057 | .054 | 97.7 | 89.4 | .009 | .065 | .058 | 97.4 | 93.9 |
| | 0.7 | $\rho$ | .003 | .124 | .142 | 92.2 | 93.7 | .002 | .129 | .142 | 93.0 | 93.7 |
| | | $\beta_1$ | -.003 | .140 | .148 | 91.1 | 90.9 | -.003 | .143 | .148 | 92.1 | 92.1 |
| | | $\beta_2$ | -.002 | .101 | .104 | 93.0 | 91.6 | -.002 | .104 | .104 | 93.5 | 93.9 |
| | | $\psi_1^2$ | -.015 | .110 | .112 | 94.8 | 86.8 | .008 | .124 | .120 | 96.2 | 90.7 |
| | | $\psi_2^2$ | -.003 | .056 | .054 | 97.7 | 89.9 | .009 | .063 | .058 | 97.8 | 93.9 |
| 10 | 0.1 | $\rho$ | .022 | .276 | .340 | 87.8 | 89.6 | .022 | .292 | .340 | 90.7 | 90.3 |
| | | $\beta_1$ | .001 | .171 | .185 | 90.6 | 89.5 | .001 | .179 | .185 | 92.3 | 92.9 |
| | | $\beta_2$ | .002 | .128 | .130 | 92.9 | 91.4 | .002 | .134 | .130 | 94.9 | 94.9 |
| | | $\psi_1^2$ | -.021 | .136 | .137 | 97.6 | 84.8 | .015 | .161 | .152 | 97.7 | 93.1 |
| | | $\psi_2^2$ | .003 | .074 | .068 | 98.6 | 91.3 | .024 | .090 | .076 | 98.2 | 96.7 |
| | 0.4 | $\rho$ | .035 | .233 | .280 | 89.6 | 89.6 | .034 | .246 | .279 | 91.7 | 91.1 |
| | | $\beta_1$ | -.002 | .171 | .186 | 90.2 | 88.6 | -.002 | .178 | .185 | 91.4 | 91.8 |
| | | $\beta_2$ | -.001 | .129 | .130 | 93.3 | 91.7 | -.001 | .134 | .130 | 94.6 | 94.7 |
| | | $\psi_1^2$ | -.019 | .136 | .140 | 97.4 | 85.7 | .017 | .161 | .156 | 97.7 | 91.9 |
| | | $\psi_2^2$ | .005 | .075 | .070 | 98.0 | 90.5 | .026 | .091 | .080 | 97.9 | 96.1 |
| | 0.7 | $\rho$ | .023 | .142 | .161 | 91.2 | 90.8 | .021 | .151 | .161 | 93.4 | 92.3 |
| | | $\beta_1$ | -.004 | .171 | .183 | 90.6 | 90.2 | -.004 | .178 | .182 | 92.2 | 92.5 |
| | | $\beta_2$ | -.004 | .126 | .130 | 93.1 | 92.4 | -.004 | .131 | .129 | 93.4 | 93.9 |
| | | $\psi_1^2$ | -.010 | .136 | .138 | 96.2 | 88.6 | .026 | .162 | .154 | 97.3 | 92.8 |
| | | $\psi_2^2$ | .004 | .073 | .069 | 97.9 | 92.6 | .025 | .088 | .077 | 97.5 | 95.4 |
| 6 | 0.1 | $\rho$ | .019 | .333 | .428 | 86.5 | 87.7 | .019 | .367 | .426 | 90.8 | 90.9 |
| | | $\beta_1$ | -.010 | .215 | .229 | 91.6 | 89.2 | -.010 | .232 | .228 | 93.9 | 93.7 |
| | | $\beta_2$ | -.006 | .171 | .173 | 92.9 | 90.9 | -.007 | .184 | .173 | 94.3 | 95.1 |
| | | $\psi_1^2$ | -.039 | .165 | .131 | 98.8 | 88.2 | .022 | .221 | .158 | 99.7 | 95.6 |
| | | $\psi_2^2$ | .015 | .102 | .082 | 99.1 | 95.1 | .054 | .139 | .101 | 99.0 | 98.4 |
| | 0.4 | $\rho$ | .024 | .286 | .362 | 88.4 | 86.6 | .024 | .314 | .359 | 91.0 | 90.1 |
| | | $\beta_1$ | -.009 | .215 | .234 | 91.1 | 88.3 | -.009 | .231 | .232 | 92.6 | 93.4 |
| | | $\beta_2$ | -.004 | .171 | .169 | 92.2 | 91.0 | -.004 | .183 | .169 | 94.0 | 94.5 |
| | | $\psi_1^2$ | -.039 | .164 | .129 | 98.7 | 90.4 | .023 | .221 | .155 | 99.6 | 96.7 |
| | | $\psi_2^2$ | .014 | .102 | .083 | 98.9 | 96.2 | .054 | .139 | .102 | 98.1 | 98.7 |
| | 0.7 | $\rho$ | .026 | .181 | .194 | 93.5 | 88.5 | .023 | .200 | .192 | 96.5 | 93.6 |
| | | $\beta_1$ | -.005 | .218 | .233 | 91.9 | 90.1 | -.005 | .233 | .231 | 93.6 | 94.8 |
| | | $\beta_2$ | -.004 | .169 | .161 | 94.0 | 92.3 | -.004 | .180 | .160 | 94.7 | 96.8 |
| | | $\psi_1^2$ | -.027 | .170 | .129 | 98.3 | 93.7 | .036 | .228 | .154 | 99.4 | 97.1 |
| | | $\psi_2^2$ | .012 | .099 | .075 | 99.7 | 97.0 | .050 | .135 | .092 | 99.3 | 99.1 |

For the ML estimate of *ρ*, bias is small and consistent across various values of *ρ* and decreases as *n* increases, ranging from -0.003 to 0.035. Both SE_*th*_ and SE_*em*_ decrease with increasing *ρ* and *n*. Overall, SE_*th*_ is smaller than SE_*em*_, with the discrepancy more pronounced for smaller sample sizes (*n* = 6 and 10). When *n* increases to 15, SE_*th*_ aligns more closely with SE_*em*_. CP_*th*_ shows a slight increasing trend with *ρ*, while CP_*b*_ does not follow the same pattern. Both CP_*th*_ and CP_*b*_ are smaller than the nominal 95% level but improve with larger *n*. REML results are similar, except that SE_*th*_ is larger, coverage probabilities are generally larger, and CP_*th*_ and CP_*b*_ are more consistent.

For the ML estimate of ***β***, bias is small and stable across values of *ρ* and decreases with *n*, ranging from -0.010 to 0.002. SE_*th*_ and SE_*em*_ also show little variation with *ρ*. Both SE metrics decrease as *n* increases, with SE_*th*_ slightly smaller than SE_*em*_. CP_*th*_ and CP_*b*_ are below the 95% level but CP_*th*_ is better than CP_*b*_; both improve with larger *n*. REML results are similar, except that SE_*th*_ is larger, coverage probabilities perform better, and CP_*th*_ and CP_*b*_ are more aligned.

For the ML estimate of 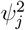, bias is small and stable across *ρ* and decreases with *n*, ranging from -0.039 to 0.014. SE_*th*_ and SE_*em*_ also show little variation with *ρ* and decrease as *n* increases. Notably, SE_*th*_ is larger than SE_*em*_ at *n* = 6. CP_*th*_ is larger than the 95% level but improves with larger *n*, while CP_*b*_ is smaller than the 95% level and has no substantial improvement with *n*. REML results are similar, except both SE metrics are larger and coverage probabilities perform better.

In summary, both ML and REML estimates exhibit small biases, and both biases and standard errors decrease with sample sizes. Coverage probabilities improve with sample sizes, except CP_*b*_ for variance components. REML generally outperforms ML in terms of coverage probability.

#### Bayesian approach

We examined several prior combinations to evaluate estimation performance. The prior for *β*_1_ and *β*_2_ were set as *N* (0, 2.5^2^), and four prior combinations were considered for variance and correlation parameters:

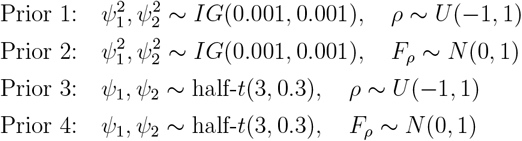

Posterior sampling was performed using the HMC method, drawing 4,000 samples for each parameter.

Table 2 presents bias and empirical coverage (EC) for each parameter. Across 4 prior combinations, bias in the *ρ* estimate increases as *ρ* increases. EC for *ρ* is close to 95% when *ρ* = 0.1 and 0.4, but slightly below 95% when *ρ* = 0.7. Bias decreases and EC improves as *n* increases. Comparing different priors on *ρ*, the Normal prior (Priors 2 and 4) slightly reduces bias and improves EC compared to the *U* (−1, 1) prior (Priors 1 and 3).

**Table 2.** Simulation results under the Bayesian framework across different numbers of trials (*n*) and true correlations (*ρ*_true_). Priors 1–4 correspond to different combinations of variance priors, either IG(0.001, 0.001) or half-*t*(3, 0.5); and correlation priors, either *U* (−1, 1) or *F*_*ρ*_ ∼ *N* (0, 1).

| $n$ | $\rho_{\text{true}}$ | Par | Prior 1 | | Prior 2 | | Prior 3 | | Prior 4 | |
| --- | --- | --- | --- | --- | --- | --- | --- | --- | --- | --- |
|  |  |  | Bias | EC | Bias | EC | Bias | EC | Bias | EC |
| 15 | 0.1 | $\rho$ | -.011 | 96.2 | -.007 | 95.5 | -.013 | 96.0 | -.009 | 95.5 |
| | | $\beta_1$ | -.003 | 94.7 | -.004 | 94.7 | -.003 | 94.1 | -.003 | 94.2 |
| | | $\beta_2$ | -.001 | 94.2 | -.001 | 94.0 | -.001 | 93.9 | -.001 | 94.6 |
| | | $\psi_1^2$ | .051 | 93.9 | .054 | 94.0 | .012 | 94.8 | .014 | 94.5 |
| | | $\psi_2^2$ | .032 | 93.4 | .033 | 93.0 | .022 | 94.5 | .023 | 94.8 |
| | 0.4 | $\rho$ | -.087 | 95.9 | -.075 | 95.7 | -.093 | 95.7 | -.081 | 95.6 |
| | | $\beta_1$ | -.008 | 94.8 | -.008 | 95.0 | -.008 | 94.2 | -.008 | 94.3 |
| | | $\beta_2$ | -.001 | 93.4 | -.001 | 93.4 | -.001 | 93.0 | -.001 | 93.3 |
| | | $\psi_1^2$ | .042 | 94.4 | .045 | 94.2 | .006 | 94.6 | .008 | 94.7 |
| | | $\psi_2^2$ | .019 | 93.7 | .021 | 93.7 | .011 | 95.1 | .012 | 95.1 |
| | 0.7 | $\rho$ | -.111 | 92.5 | -.094 | 93.6 | -.117 | 91.2 | -.101 | 93.0 |
| | | $\beta_1$ | .006 | 95.0 | .006 | 95.0 | .006 | 94.7 | .006 | 94.6 |
| | | $\beta_2$ | .003 | 93.5 | .003 | 94.1 | .003 | 93.0 | .004 | 93.7 |
| | | $\psi_1^2$ | .028 | 93.9 | .033 | 93.8 | -.004 | 93.3 | .000 | 93.7 |
| | | $\psi_2^2$ | .011 | 92.6 | .014 | 92.7 | .002 | 94.0 | .004 | 94.6 |
| 10 | 0.1 | $\rho$ | -.024 | 96.6 | -.019 | 96.0 | -.025 | 96.4 | -.021 | 94.9 |
| | | $\beta_1$ | -.006 | 94.0 | -.007 | 94.2 | -.006 | 92.7 | -.006 | 93.3 |
| | | $\beta_2$ | .003 | 92.5 | .003 | 92.6 | .003 | 92.3 | .003 | 92.1 |
| | | $\psi_1^2$ | .088 | 90.9 | .093 | 91.1 | .017 | 90.8 | .020 | 91.3 |
| | | $\psi_2^2$ | .044 | 94.3 | .047 | 94.5 | .024 | 96.3 | .025 | 96.1 |
| | 0.4 | $\rho$ | -.118 | 95.0 | -.102 | 94.9 | -.122 | 94.4 | -.108 | 94.7 |
| | | $\beta_1$ | .002 | 93.3 | .002 | 93.4 | .002 | 91.8 | .002 | 92.1 |
| | | $\beta_2$ | .010 | 93.7 | .009 | 94.3 | .010 | 94.1 | .009 | 94.3 |
| | | $\psi_1^2$ | .074 | 92.3 | .080 | 92.1 | .009 | 92.7 | .012 | 92.7 |
| | | $\psi_2^2$ | .040 | 94.6 | .043 | 94.3 | .020 | 96.6 | .022 | 97.0 |
| | 0.7 | $\rho$ | -.159 | 92.2 | -.137 | 93.7 | -.165 | 91.1 | -.143 | 92.7 |
| | | $\beta_1$ | -.006 | 91.8 | -.006 | 91.4 | -.006 | 90.8 | -.006 | 90.9 |
| | | $\beta_2$ | -.006 | 92.4 | -.006 | 93.2 | -.006 | 92.5 | -.006 | 92.9 |
| | | $\psi_1^2$ | .031 | 92.3 | .038 | 92.7 | -.018 | 92.2 | -.013 | 92.1 |
| | | $\psi_2^2$ | .015 | 92.7 | .019 | 93.1 | .000 | 95.0 | .002 | 95.2 |
| 6 | 0.1 | $\rho$ | -.025 | 97.9 | -.019 | 96.5 | -.026 | 96.7 | -.020 | 95.8 |
| | | $\beta_1$ | -.010 | 92.0 | -.010 | 92.1 | -.008 | 90.3 | -.008 | 89.8 |
| | | $\beta_2$ | .011 | 94.9 | .011 | 94.9 | .011 | 94.4 | .011 | 94.2 |
| | | $\psi_1^2$ | .205 | 92.5 | .217 | 92.4 | .025 | 92.8 | .030 | 92.5 |
| | | $\psi_2^2$ | .104 | 96.4 | .109 | 96.2 | .037 | 97.8 | .040 | 97.8 |
| | 0.4 | $\rho$ | -.156 | 95.9 | -.136 | 96.0 | -.156 | 94.3 | -.139 | 95.0 |
| | | $\beta_1$ | -.003 | 94.5 | -.004 | 94.6 | -.001 | 93.0 | -.001 | 93.1 |
| | | $\beta_2$ | .004 | 95.3 | .003 | 95.6 | .004 | 95.1 | .004 | 95.0 |
| | | $\psi_1^2$ | .161 | 93.9 | .173 | 94.1 | .009 | 93.7 | .015 | 94.0 |
| | | $\psi_2^2$ | .080 | 97.3 | .085 | 96.8 | .024 | 98.5 | .026 | 98.5 |
| | 0.7 | $\rho$ | -.239 | 93.2 | -.207 | 94.3 | -.239 | 91.8 | -.210 | 93.5 |
| | | $\beta_1$ | -.001 | 94.9 | -.001 | 94.5 | .000 | 93.0 | .000 | 93.3 |
| | | $\beta_2$ | .008 | 93.2 | .008 | 92.6 | .008 | 92.9 | .008 | 92.9 |
| | | $\psi_1^2$ | .117 | 94.7 | .128 | 94.8 | -.007 | 94.4 | -.002 | 94.4 |
| | | $\psi_2^2$ | .051 | 97.3 | .056 | 97.5 | .009 | 98.6 | .010 | 98.4 |

For 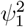 and 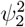, bias decreases as *ρ* increases, and EC is close to 95% although it is less stable for small *n* = 6 and 10. Bias decreases and EC improves with larger *n*. Among all priors on 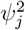, the half-*t*(3, 0.3) prior (Priors 3 and 4) yields smaller bias and better EC than the *IG*(0.001, 0.001) prior (Priors 1 and 2), particularly for small sample sizes. For *β*_1_ and *β*_2_, estimates exhibit small biases and reasonable ECs, which are not largely affected by priors on *ρ*, 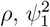, and 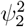. Overall, Prior 4 demonstrates the most robust performance across all parameters.

Compared to the frequentist approach, Bayesian estimates show slightly larger biases, likely due to the use of flat or weakly informative priors. Nevertheless, EC is closer to the nominal level of 0.95, indicating that the Bayesian approach provides more reliable interval estimates.

### 3.2 Real Application

To demonstrate the proposed method in a real-world setting, we analyzed data from 14 non–small cell lung cancer (NSCLC) trials comparing chemotherapy and immunotherapy [15–27]. We considered two scenarios: the first evaluated the T-association between progression-free survival (PFS) and OS, and the second evaluated the T-association between objective response and OS. Based on the simulation results, the REML approach was selected for the frequentist analysis, and Prior 4 was used for the Bayesian analysis, as it demonstrated the most robust performance across parameters.

#### Scenario 1

All 14 NSCLC trials reported hazard ratios for PFS and OS, which were used to examine the T-association between these endpoints. Figure 2(a) visualizes the log(HR) estimates for PFS and OS across the 14 trials. The half-widths and half-heights of the ellipses represent 1.96 times the within-trial standard errors.

**Figure 2.**
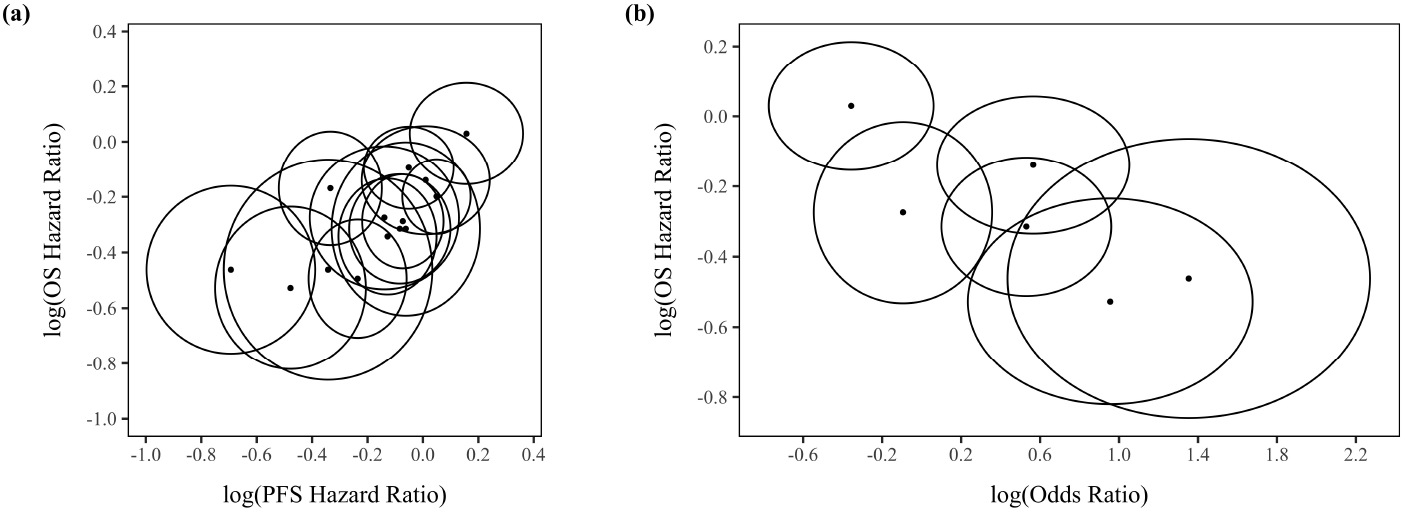
Visualization of transformed treatment effect estimates (solid points) for the two scenarios. The half-widths and half-heights of the ellipses represent 1.96 times the within-trial standard errors. (a) log(HR) for OS versus log(HR) for PFS across 14 trials in Scenario 1. (b) log(HR) for OS versus log(OR) for objective response across 6 trials in Scenario 2.

Using the proposed method, the estimated correlation (*ρ*) between the log(HR) estimates on PFS and OS was 0.71 (95% bootstrapped CI: 0.36 to 0.91; 95% normal-based CI: 0.32 to 0.90) using the REML approach; and 0.63 (95% CrI: 0.23 to 0.88) using the Bayesian approach (Table 3). These results show a strong correlation between treatment effects on PFS and OS, offering supportive evidence for FDA consideration regarding the validity of PFS as a surrogate endpoint for OS in terms of T-association. For a reference, the Spearman correlation, which ignores within-study variability, was 0.76 (95% bootstrapped CI: 0.33 to 0.95), showing a higher correlation estimate and a wider interval than REML. This wider interval is probably caused by the ignorance of within-trial variances.

**Table 3.** Estimates of the model parameters with 95% confidence/credible intervals for two real-world scenarios.

| Scenario | Par | REML |  |  | Bayesian |  |
| --- | --- | --- | --- | --- | --- | --- |
|  |  | Estimates | 95% Normal-based CI | 95% bootstrapped CI | Posterior mean | 95% CrI |
| 1 | $\rho$ | 0.71 | (0.32, 0.90) | (0.36, 0.91) | 0.63 | (0.23, 0.88) |
| | $\beta_1$ | -0.15 | (-0.25, -0.04) | (-0.25, -0.04) | -0.14 | (-0.27, -0.03) |
| | $\beta_2$ | -0.26 | (-0.35, -0.18) | (-0.34, -0.18) | -0.26 | (-0.36, -0.17) |
| | $\psi_1^2$ | 0.03 | (0.009, 0.10) | (0.005, 0.07) | 0.04 | (0.009, 0.10) |
| | $\psi_2^2$ | 0.01 | (0.003, 0.06) | (0.001, 0.04) | 0.02 | (0.002, 0.05) |
| 2 | $\rho$ | -0.79 | (-0.96, -0.16) | (-0.98, -0.16) | -0.56 | (-0.90, 0.07) |
| | $\beta_1$ | 0.40 | (-0.09, 0.90) | (-0.08, 0.88) | 0.38 | (-0.09, 0.91) |
| | $\beta_2$ | -0.24 | (-0.42, -0.08) | (-0.41, -0.08) | -0.24 | (-0.45, -0.06) |
| | $\psi_1^2$ | 0.29 | (0.05, 1.63) | (0.03, 0.94) | 0.27 | (0.03, 0.91) |
| | $\psi_2^2$ | 0.03 | (0.004, 0.19) | (0.002, 0.10) | 0.04 | (0.001, 0.14) |

#### Scenario 2

Among the 14 NSCLC trials, 6 trials reported odds ratios for objective response comparing chemotherapy and immunotherapy, allowing assessment of the T-association between objective response and OS. Objective response, defined as a complete or partial tumor response, is a potential surrogate endpoint that can be observed earlier than OS. Figure 2(b) displays the transformed treatment effect estimates from the six trials.

Using the proposed method, the estimated correlation between treatment effects on objective response and OS was -0.79 (95% bootstrapped CI: -0.98 to -0.16; 95% normal-based CI: -0.96 to -0.16) using the REML approach (Table 3). The Bayesian approach also showed a negative correlation of -0.56, but with a wider and more conservative credible interval (95% CrI: -0.90 to -0.07). These results provide evidence that the objective response may serve as a surrogate endpoint for OS. For a reference, the Spearman correlation was -0.77 (bootstrapped CI unavailable due to the limited number of trials).

## 4 Discussion

According to FDA guidelines, a biomarker is considered a valid surrogate if it satisfies two key criteria: I-association and T-association. I-association is commonly evaluated, but T-association is often overlooked, likely due to the absence of suitable analysis methods. To fill this gap, we propose a new analysis method based on the BRMA model to evaluate T-association. Parameter estimation is performed using both frequentist and Bayesian approaches. We believe the proposed method will serve as the statistical foundation for future FDA Accelerated Approval drugs.

Our simulation results indicate that, compared to REML, the ML approach is less stable, sometimes yielding narrower CIs and smaller coverage probability for the key correlation. Therefore, when the number of studies is limited, the REML approach is preferable. In addition, for small sample sizes, large-sample normal-based coverage probabilities for variance components under both ML and REML tend to be overly large due to large standard errors and wide CIs. When fewer than 10 studies are available, we recommend constructing CIs for variance components using the parametric bootstrap method under REML when applying a frequentist approach.

Compared to the frequentist approach, the Bayesian approach provides more reliable interval estimates although its point estimates may have larger bias due to the influence of priors. Simulation results suggest that the half-*t*(*ν, τ*) prior on *ψ*_*j*_ is more robust than the inverse-gamma *IG*(*a, b*) prior on 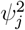, and the Normal prior on the Fisher-*z* transformed correlation is preferable to the uniform *U* (−1, 1) prior on *ρ*. Specifying a prior on the Fisher-*z* transformed correlation allows sampling on the unconstrained real line rather than within a bounded interval, which improves numerical stability and MCMC efficiency, as suggested in reference [14].

The number of trials available to evaluate T-association is often limited, raising the question of the minimum number of trials required to obtain reliable correlation estimates. Based on our simulation results, we recommend including at least six trials. Under the frequentist framework, the ML method estimates five parameters, while the REML method estimates three. When the true correlation is moderate (e.g., *ρ* = 0.4), using only five trials led to a high proportion of divergent estimates across 1,000 simulations: 57.4% for ML and 44% for REML. Although the Bayesian approach does not encounter such numerical divergence, its performance at very small sample sizes is also unstable.

The bivariate normal assumption in the BRMA model may be restrictive, although the observed treatment effects on the biomarker of interest and the true endpoint are generally assumed to individually follow a normal distribution. To assess the validity of this assumption, we introduce a simple diagnostic procedure: applying the Shapiro–Wilk test to check whether the standardized residuals 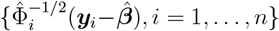 approximate the standard normal distribution. In the real-data application, the Shapiro–Wilk test on the standardized residuals yielded *p*-values of 0.25 and 0.30 for the two scenarios, providing no evidence against normality.

Clinical trial designs can differ in various aspects, such as treatment regimen, leading to heterogeneity across studies. The proposed method can theoretically be extended to account for such heterogeneity by incorporating trial-level covariates. Specifically, the parameter ***β*** in the BRMA model can be replaced with 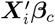, where ***X***_*i*_ is the covariate matrix for study *i*, and ***β***_*c*_ is the corresponding coefficient vector. However, due to the limited number of trials available in practice, we do not perform this extension in the current study.

## Data Availability

All data produced in the present work are contained in the manuscript

## Supplementary Material

1. Online supplemental material.
2. The freely available R package at https://github.com/jybelindahung/T-association.

