## Supplemental Materials for "Evaluation Methods for T-association of a Surrogate Endpoint"

### Appendix A

#### *Maximum likelihood*

As specified in Section 2.2, the parameters include $\boldsymbol{\theta} = (\boldsymbol{\beta}', \boldsymbol{\psi}', \rho)$, and the corresponding log-likelihood function is:

$$\ell\left(\boldsymbol{\theta}|\boldsymbol{y}, \boldsymbol{s}\right) = -\frac{1}{2}\Big[n\log(2\pi) + \log|\boldsymbol{\Phi}| + (\boldsymbol{y} - \boldsymbol{X}\boldsymbol{\beta})'\boldsymbol{\Phi}^{-1}(\boldsymbol{y} - \boldsymbol{X}\boldsymbol{\beta})\Big],$$

The components of the score vector $\boldsymbol{U}(\boldsymbol{\theta}) = \Big(\boldsymbol{U}_{\boldsymbol{\beta}}(\boldsymbol{\theta})', \boldsymbol{U}_{\psi_1^2}(\boldsymbol{\theta}), \boldsymbol{U}_{\psi_2^2}(\boldsymbol{\theta}), U_\rho(\boldsymbol{\theta})\Big)'$ are given by:

$$\boldsymbol{U}_{\boldsymbol{\beta}}(\boldsymbol{\theta}) = \sum_{i=1}^{n} \boldsymbol{X}_i'\Phi_i^{-1}(\boldsymbol{y}_i - \boldsymbol{X}_i\boldsymbol{\beta}),$$

$$U_{\psi_j^2}(\boldsymbol{\theta}) = \sum_{i=1}^{n}\left[-\frac{1}{2}\operatorname{tr}\Big(\Phi_i^{-1}\frac{\partial\Phi_i}{\partial\psi_j^2}\Big) + \frac{1}{2}(\boldsymbol{y}_i - \boldsymbol{X}_i\boldsymbol{\beta})'\Phi_i^{-1}\frac{\partial\Phi_i}{\partial\psi_j^2}\Phi_i^{-1}(\boldsymbol{y}_i - \boldsymbol{X}_i\boldsymbol{\beta})\right], \quad j = 1, 2,$$

$$U_\rho(\boldsymbol{\theta}) = \sum_{i=1}^{n}\left[-\frac{1}{2}\operatorname{tr}\Big(\Phi_i^{-1}\frac{\partial\Phi_i}{\partial\rho}\Big) + \frac{1}{2}(\boldsymbol{y}_i - \boldsymbol{X}_i\boldsymbol{\beta})'\Phi_i^{-1}\frac{\partial\Phi_i}{\partial\rho}\Phi_i^{-1}(\boldsymbol{y}_i - \boldsymbol{X}_i\boldsymbol{\beta})\right].$$

The first order derivatives of $\Phi_i$ with respect to each of the parameters are:

$$\frac{\partial\Phi_i}{\partial\psi_1^2} = \begin{pmatrix} 1 & \dfrac{\rho(\psi_2^2 + s_{i2}^2)}{2\sqrt{(\psi_1^2 + s_{i1}^2)(\psi_2^2 + s_{i2}^2)}} \\ \dfrac{\rho(\psi_2^2 + s_{i2}^2)}{2\sqrt{(\psi_1^2 + s_{i1}^2)(\psi_2^2 + s_{i2}^2)}} & 0 \end{pmatrix},$$

$$\frac{\partial\Phi_i}{\partial\psi_2^2} = \begin{pmatrix} 0 & \dfrac{\rho(\psi_1^2 + s_{i1}^2)}{2\sqrt{(\psi_1^2 + s_{i1}^2)(\psi_2^2 + s_{i2}^2)}} \\ \dfrac{\rho(\psi_1^2 + s_{i1}^2)}{2\sqrt{(\psi_1^2 + s_{i1}^2)(\psi_2^2 + s_{i2}^2)}} & 1 \end{pmatrix},$$

$$\frac{\partial\Phi_i}{\partial\rho} = \begin{pmatrix} 0 & \sqrt{(\psi_1^2 + s_{i1}^2)(\psi_2^2 + s_{i2}^2)} \\ \sqrt{(\psi_1^2 + s_{i1}^2)(\psi_2^2 + s_{i2}^2)} & 0 \end{pmatrix}.$$

The MLE is obtained by solving $\boldsymbol{U}(\boldsymbol{\theta}) = \boldsymbol{0}$ with the Newton-Raphson algorithm.

The asymptotic variance are derived by the inverse of the observed information matrix $\boldsymbol{\mathcal{I}}^{-1}$, where $\boldsymbol{\mathcal{I}}$ is defined by:

$$\boldsymbol{\mathcal{I}} = -\left.\frac{\partial^2 \ell(\boldsymbol{\theta} \mid \boldsymbol{y}, \boldsymbol{s})}{\partial \boldsymbol{\theta}\, \partial \boldsymbol{\theta}'}\right|_{\boldsymbol{\theta}=\hat{\boldsymbol{\theta}}}$$

and each of the components in the matrix is show below:

$$\frac{\partial^2 \ell(\boldsymbol{\theta} \mid \boldsymbol{y}, \boldsymbol{s})}{\partial \boldsymbol{\beta}\, \partial \boldsymbol{\beta}'} = -\sum_{i=1}^{n} \boldsymbol{X}_i' \Phi_i^{-1} \boldsymbol{X}_i,$$

$$\frac{\partial^2 \ell(\boldsymbol{\theta} \mid \boldsymbol{y}, \boldsymbol{s})}{\partial \boldsymbol{\beta}\, \partial \psi_j^2} = -\sum_{i=1}^{n} \boldsymbol{X}_i' \Phi_i^{-1} \frac{\partial \Phi_i}{\partial \psi_j^2} \Phi_i^{-1} (\boldsymbol{y}_i - \boldsymbol{X}_i \boldsymbol{\beta}), \quad j = 1, 2,$$

$$\frac{\partial^2 \ell(\boldsymbol{\theta} \mid \boldsymbol{y}, \boldsymbol{s})}{\partial \boldsymbol{\beta}\, \partial \rho} = -\sum_{i=1}^{n} \boldsymbol{X}_i' \Phi_i^{-1} \frac{\partial \Phi_i}{\partial \rho} \Phi_i^{-1} (\boldsymbol{y}_i - \boldsymbol{X}_i \boldsymbol{\beta}).$$

For the second derivatives with respect to the variance and correlation parameters $\theta_a, \theta_b \in \{\psi_1^2, \psi_2^2, \rho\}$, they share the formula:

$$\begin{aligned} \frac{\partial^2 \ell(\boldsymbol{\theta} \mid \boldsymbol{y}, \boldsymbol{s})}{\partial \theta_a\, \partial \theta_b} &= \sum_{i=1}^{n} \frac{1}{2} \Bigg[ \mathrm{tr}\Big( \Phi_i^{-1} \frac{\partial \Phi_i}{\partial \theta_a} \Phi_i^{-1} \frac{\partial \Phi_i}{\partial \theta_b} \Big) - \mathrm{tr}\Big( \Phi_i^{-1} \frac{\partial^2 \Phi_i}{\partial \theta_a \partial \theta_b} \Big) \\ &\quad + (\boldsymbol{y}_i - \boldsymbol{X}_i \boldsymbol{\beta})' \Big( \Phi_i^{-1} \frac{\partial^2 \Phi_i}{\partial \theta_a \partial \theta_b} \Phi_i^{-1} - \Phi_i^{-1} \frac{\partial \Phi_i}{\partial \theta_a} \Phi_i^{-1} \frac{\partial \Phi_i}{\partial \theta_b} \Phi_i^{-1} - \Phi_i^{-1} \frac{\partial \Phi_i}{\partial \theta_b} \Phi_i^{-1} \frac{\partial \Phi_i}{\partial \theta_a} \Phi_i^{-1} \Big) (\boldsymbol{y}_i - \boldsymbol{X}_i \boldsymbol{\beta}) \Bigg]. \end{aligned}$$

The second order derivatives of $\Phi_i$ are:

$$\frac{\partial^2 \Phi_i}{\partial (\psi_1^2)^2} = \begin{pmatrix} 0 & -\dfrac{\rho(\psi_2^2 + s_{i2}^2)}{4(\psi_1^2 + s_{i1}^2)^{3/2}(\psi_2^2 + s_{i2}^2)^{1/2}} \\ -\dfrac{\rho(\psi_2^2 + s_{i2}^2)}{4(\psi_1^2 + s_{i1}^2)^{3/2}(\psi_2^2 + s_{i2}^2)^{1/2}} & 0 \end{pmatrix},$$

$$\frac{\partial^2 \Phi_i}{\partial (\psi_2^2)^2} = \begin{pmatrix} 0 & -\dfrac{\rho(\psi_1^2 + s_{i1}^2)}{4(\psi_1^2 + s_{i1}^2)^{1/2}(\psi_2^2 + s_{i2}^2)^{3/2}} \\ -\dfrac{\rho(\psi_1^2 + s_{i1}^2)}{4(\psi_1^2 + s_{i1}^2)^{1/2}(\psi_2^2 + s_{i2}^2)^{3/2}} & 0 \end{pmatrix},$$

$$\frac{\partial^2 \Phi_i}{\partial \rho^2} = \begin{pmatrix} 0 & 0 \\ 0 & 0 \end{pmatrix},$$

$$\frac{\partial^2 \Phi_i}{\partial \psi_1^2 \partial \psi_2^2} = \begin{pmatrix} 0 & -\dfrac{\rho}{4\sqrt{(\psi_1^2 + s_{i1}^2)(\psi_2^2 + s_{i2}^2)}} \\ -\dfrac{\rho}{4\sqrt{(\psi_1^2 + s_{i1}^2)(\psi_2^2 + s_{i2}^2)}} & 0 \end{pmatrix},$$

$$\frac{\partial^2 \Phi_i}{\partial \psi_1^2 \partial \rho} = \begin{pmatrix} 0 & \dfrac{(\psi_2^2 + s_{i2}^2)}{2\sqrt{(\psi_1^2 + s_{i1}^2)(\psi_2^2 + s_{i2}^2)}} \\ \dfrac{(\psi_2^2 + s_{i2}^2)}{2\sqrt{(\psi_1^2 + s_{i1}^2)(\psi_2^2 + s_{i2}^2)}} & 0 \end{pmatrix},$$

$$\frac{\partial^2 \Phi_i}{\partial \psi_2^2 \partial \rho} = \begin{pmatrix} 0 & \dfrac{(\psi_1^2 + s_{i1}^2)}{2\sqrt{(\psi_1^2 + s_{i1}^2)(\psi_2^2 + s_{i2}^2)}} \\ \dfrac{(\psi_1^2 + s_{i1}^2)}{2\sqrt{(\psi_1^2 + s_{i1}^2)(\psi_2^2 + s_{i2}^2)}} & 0 \end{pmatrix}.$$

##### *Restricted maximum likelihood*

For the restricted likelihood approach, as specified in Section 2.2, the parameters include $\boldsymbol{\theta}_R = (\psi_1^2, \psi_2^2, \rho)$, and the corresponding log-likelihood function is

$$\ell_R\left(\boldsymbol{\theta}_R|\boldsymbol{y}, \boldsymbol{s}\right) \propto -\frac{1}{2}\Big[\log|\boldsymbol{\Phi}| + \log\left|\boldsymbol{X}'\boldsymbol{\Phi}^{-1}\boldsymbol{X}\right| + (\boldsymbol{y} - \boldsymbol{X}\boldsymbol{\beta}_R)'\boldsymbol{\Phi}^{-1}(\boldsymbol{y} - \boldsymbol{X}\boldsymbol{\beta}_R)\Big],$$

where $\boldsymbol{\beta}_R = (\boldsymbol{X}'\boldsymbol{\Phi}^{-1}\boldsymbol{X})^{-1}\boldsymbol{X}'\boldsymbol{\Phi}^{-1}\boldsymbol{y} = (\sum_{i=1}^n \boldsymbol{X}_i'\Phi_i^{-1}\boldsymbol{X}_i)^{-1}(\sum_{i=1}^n \boldsymbol{X}_i'\Phi_i^{-1}\boldsymbol{y}_i)$. The score functions with respect to each parameter $\theta \in \{\psi_1^2, \psi_2^2, \rho\}$ are

$$\begin{aligned}\frac{\partial \ell_R\left(\boldsymbol{\theta}_R|\boldsymbol{y}, \boldsymbol{s}\right)}{\partial\theta} = -\frac{1}{2}\sum_{i=1}^n \Bigg[&\operatorname{tr}\Big(\Phi_i^{-1}\frac{\partial\Phi_i}{\partial\theta}\Big) - \operatorname{tr}\Bigg(\Big(\sum_{j=1}^n \boldsymbol{X}_j'\Phi_j^{-1}\boldsymbol{X}_j\Big)^{-1}\boldsymbol{X}_i'\Phi_i^{-1}\frac{\partial\Phi_i}{\partial\theta}\Phi_i^{-1}\boldsymbol{X}_i\Bigg) \\ &- (\boldsymbol{y}_i - \boldsymbol{X}_i\boldsymbol{\beta}_R)'\Phi_i^{-1}\frac{\partial\Phi_i}{\partial\theta}\Phi_i^{-1}(\boldsymbol{y}_i - \boldsymbol{X}_i\boldsymbol{\beta}_R)\Bigg].\end{aligned}$$

The REML estimates for $\boldsymbol{\theta}_R$ are obtained by solving $\boldsymbol{U}_R(\boldsymbol{\theta}_R) = \boldsymbol{0}$ numerically using the Newton-Raphson algorithm. Once $\hat{\boldsymbol{\theta}}_R$ is obtained, it is plugged into $\boldsymbol{\beta}_R = (\boldsymbol{X}'\boldsymbol{\Phi}^{-1}\boldsymbol{X})^{-1}\boldsymbol{X}'\boldsymbol{\Phi}^{-1}\boldsymbol{y}$ to compute the corresponding estimator for $\boldsymbol{\beta}$.

The variance of the mean effects $\boldsymbol{\beta}$ are estimated by $(\boldsymbol{X}'\hat{\boldsymbol{\Phi}}^{-1}\boldsymbol{X})^{-1}$. For the covariance components (variances and correlation), the asymptotic variances are obtained from the inverse of the observed information matrix. The observed information matrix for $\boldsymbol{\theta}_R$ is defined by:

$$\boldsymbol{\mathcal{I}}_R = -\left.\frac{\partial^2\ell_R(\boldsymbol{\theta}_R \mid \boldsymbol{y}, \boldsymbol{s})}{\partial\boldsymbol{\theta}_R\,\partial\boldsymbol{\theta}_R'}\right|_{\boldsymbol{\theta}_R = \hat{\boldsymbol{\theta}}_R}$$

and for $\theta_a, \theta_b \in \{\psi_1^2, \psi_2^2, \rho\}$, the explicit formula is:

$$\begin{aligned}\frac{\partial^2\ell_R(\boldsymbol{\theta}_R \mid \boldsymbol{y}, \boldsymbol{s})}{\partial\theta_a\,\partial\theta_b} = -\frac{1}{2}\sum_{i=1}^n \Bigg[\ &\operatorname{tr}\Big(\Phi_i^{-1}\frac{\partial^2\Phi_i}{\partial\theta_a\partial\theta_b}\Big) - \operatorname{tr}\Big(\Phi_i^{-1}\frac{\partial\Phi_i}{\partial\theta_a}\Phi_i^{-1}\frac{\partial\Phi_i}{\partial\theta_b}\Big) \\ - \operatorname{tr}&\Bigg(\Big(\sum_{j=1}^n \boldsymbol{X}_j^\top\Phi_j^{-1}\boldsymbol{X}_j\Big)^{-1}\Big(\sum_{j=1}^n \boldsymbol{X}_j^\top\Phi_j^{-1}\frac{\partial\Phi_j}{\partial\theta_a}\Phi_j^{-1}\boldsymbol{X}_j\Big)\Big(\sum_{j=1}^n \boldsymbol{X}_j^\top\Phi_j^{-1}\boldsymbol{X}_j\Big)^{-1}\boldsymbol{X}_i^\top\Phi_i^{-1}\,\frac{\partial\Phi_i}{\partial\theta_b}\,\Phi_i^{-1}\boldsymbol{X}_i\Bigg) \\ - \operatorname{tr}&\Bigg(\Big(\sum_{j=1}^n \boldsymbol{X}_j^\top\Phi_j^{-1}\boldsymbol{X}_j\Big)^{-1}\boldsymbol{X}_i^\top\Phi_i^{-1}\,\frac{\partial^2\Phi_i}{\partial\theta_a\partial\theta_b}\,\Phi_i^{-1}\boldsymbol{X}_i\Bigg) \\ + \operatorname{tr}&\Bigg(\Big(\sum_{j=1}^n \boldsymbol{X}_j^\top\Phi_j^{-1}\boldsymbol{X}_j\Big)^{-1}\boldsymbol{X}_i^\top\Phi_i^{-1}\,\frac{\partial\Phi_i}{\partial\theta_a}\,\Phi_i^{-1}\frac{\partial\Phi_i}{\partial\theta_b}\,\Phi_i^{-1}\boldsymbol{X}_i\Bigg) \\ + \operatorname{tr}&\Bigg(\Big(\sum_{j=1}^n \boldsymbol{X}_j^\top\Phi_j^{-1}\boldsymbol{X}_j\Big)^{-1}\boldsymbol{X}_i^\top\Phi_i^{-1}\,\frac{\partial\Phi_i}{\partial\theta_b}\,\Phi_i^{-1}\frac{\partial\Phi_i}{\partial\theta_a}\,\Phi_i^{-1}\boldsymbol{X}_i\Bigg) \\ + 2&(\boldsymbol{y}_i - \boldsymbol{X}_i\hat{\boldsymbol{\beta}})'\Phi_i^{-1}\frac{\partial\Phi_i}{\partial\theta_b}\Phi_i^{-1}\boldsymbol{X}_i\frac{\partial\hat{\boldsymbol{\beta}}}{\partial\theta_a} \\ - (&\boldsymbol{y}_i - \boldsymbol{X}_i\hat{\boldsymbol{\beta}})'\Bigg(-\Phi_i^{-1}\frac{\partial\Phi_i}{\partial\theta_a}\Phi_i^{-1}\frac{\partial\Phi_i}{\partial\theta_b}\Phi_i^{-1} - \Phi_i^{-1}\frac{\partial\Phi_i}{\partial\theta_b}\Phi_i^{-1}\frac{\partial\Phi_i}{\partial\theta_a}\Phi_i^{-1} + \Phi_i^{-1}\frac{\partial^2\Phi_i}{\partial\theta_a\partial\theta_b}\Phi_i^{-1}\Bigg)(\boldsymbol{y}_i - \boldsymbol{X}_i\hat{\boldsymbol{\beta}})\ \Bigg],\end{aligned}$$

where

$$\frac{\partial\hat{\boldsymbol{\beta}}}{\partial\theta_a} = -\left(\sum_{j=1}^n \boldsymbol{X}_j'\Phi_j^{-1}\boldsymbol{X}_j\right)^{-1}\sum_{j=1}^n \boldsymbol{X}_j'\Phi_j^{-1}\frac{\partial\Phi_j}{\partial\theta_a}\Phi_j^{-1}(\boldsymbol{y}_j - \boldsymbol{X}_j\hat{\boldsymbol{\beta}}).$$
